# Wastewater Surveillance for SARS-CoV-2 in an Atlanta, Georgia Jail: A study of the feasibility of wastewater monitoring and correlation of building wastewater and individual testing results

**DOI:** 10.1101/2023.05.17.23290000

**Authors:** Lindsay B. Saber, Shanika Kennedy, Yixin Yang, Kyler Moore, Yuke Wang, Stephen P. Hilton, Tylis Chang, Pengbo Liu, Victoria L. Phillips, Matthew J. Akiyama, Christine L. Moe, Anne C. Spaulding

## Abstract

**Background:** Wastewater-based surveillance (WBS) on an institutional level was implemented during the COVID-19 pandemic, including carceral facilities. In this study of a mega-jail, we examined the relationship between COVID-19 diagnostic test results from jail residents and the PCR signal for SARS-CoV-2 detection in weekly samples of jail wastewater over a 28-week period.

**Methods:** This study in a Georgia Jail (average population ∼2,700) was conducted October 2021-May 2022. Weekly on-site wastewater samples were collected (Moore Swabs) and tested for SARS-CoV-2 RNA using RTqPCR. The source of wastewater was identified using a tracer dye. The jail offered residents rapid antigen testing at entry. We conducted periodic mass screenings via RT-PCR of nasal swabs. We aggregated individual test data, calculated the Spearman correlation coefficient, and performed logistic regression to examine the relationship between the strength of the SARS-CoV-2 PCR signal (Ct value) in wastewater and the proportion of the jail population that tested positive for COVID-19.

**Results:** Overall, 3770 individual nasal specimens were collected; 3.4% were COVID-positive. Weekly diagnostic test positivity ranged from 0%-29.5%. Dye tests demonstrated that a single wastewater collection point was sufficiently representative of the jail’s aggregate viral load. Twenty-five wastewater samples were collected. RT-qPCR Ct values for wastewater samples with SARS-CoV-2 RNA ranged from 28.1-39.9. A strong inverse correlation was observed between diagnostic test positivity and Ct value (r= −0.67, p < 0.01).

**Conclusion:** WBS was shown to be an effective strategy for surveilling COVID-19 in a large jail. Strong partnerships with the jail administration are essential to the success of WBS surveillance.

**Key Points:**

- Monitoring the wastewater of a large jail can be effective for infectious disease surveillance.
- To diagnose COVID-19, jail residents used self-collected nasal specimens.
- There was a strong correlation between the prevalence of COVID-19 cases and the SARS-CoV-2 PCR signal in wastewater samples from the jail.

## Introduction

Factors facilitating SARS-CoV-2 transmission include crowding, mask shortages, and insufficient quarantining and isolation[2]. Jails—short-term carceral institutions—experienced these factors when the COVID-19 pandemic started in 2020[3]. That year, 7% of US jails were over-capacity, even though total admissions dropped from 10.3 million in 2019, to 8.7 million (16%) [4]. The Centers for Disease Control and Prevention (CDC) has published specific mitigation recommendations for carceral facilities [5–7]. Nonetheless, carceral COVID-19 incidence exceeded that of surrounding communities up to five-fold [8]. To regularly screen thousands of individuals in a mega-jail for an often-asymptomatic disease poses logistic challenges, e.g., coordinating the timing of collecting tests with other activities [6, 9–13]. Wastewater-based surveillance (WBS) can detect SARS-CoV-2 at an institutional level, and if implemented in jails, could potentially save time, resources, and lives [14].

Sampling wastewater via manholes downstream of a building, holds the potential of surveilling for SARS-CoV-2 in that building’s population. WBS may detect SARS-CoV-2 prior to clinical symptoms and serve as a sensitive, low-cost, non-invasive early warning tool [15–20]. This study sought to validate WBS for monitoring SARS-CoV-2 infection in a large jail.

## Methods

### Setting and Study Population

The rated capacity of Fulton County (Georgia) Jail is 2,600 persons [21]. Mean population during our study period, October 2021 - May 2022, was 2,700 persons (SD=133). The main complex has north and south towers, each with seven floors, and six housing units per floor. People move to housing units within 24 hours after entry. Most new entrants go to one floor of the south tower that is designated to house new admissions. Housing units usually hold 40 persons maximum, in -up to 20 two-person cells. When resident volume exceeds capacity, extra mattresses in durable plastic frames (which residents call “boats”) are placed on the floor. The study population was persons in custody at the main complex of the jail, who on an average shift outnumber correctional officers over 15-fold. The jail provided demographic information on the predominately male study population at the main complex; after women are booked, most are moved to an annex located 14 miles south of the main complex. As of October 2022, 93% of the population was held on felony charges. The median length of stay has historically been 5 days [22], but this was longer during the pandemic.

### Wastewater Monitoring

An Emory University sampling team collected weekly wastewater samples from the jail through the project period. Moore swabs (see Figure 1) were suspended overnight in manhole sites around the jail property (Figure 2) [23, 24]. Eluted wastewater from the swabs was tested using real time, quantitative reverse transcription-polymerase chain reaction (RT-qPCR) at the Center for Global Safe WASH Laboratory of Emory as previously described [25, 26]. The amount of SARS-CoV-2 viral RNA present in a sample was measured by the RT-qPCR cycle threshold (Ct) value, which was inversely related to the concentration of SARS-CoV-2 in the Moore swab eluate. Positive samples were defined as those with RT-qPCR results in both duplicate wells <40 Ct and within 2 Ct of each other.

**Figure 1.**
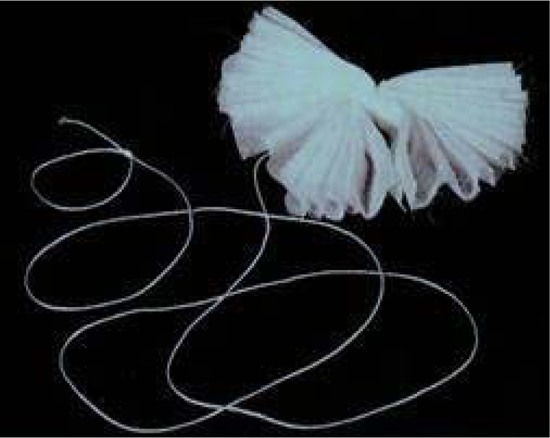
Moore Swab. 4” by 4” cotton gauze squares, tied together with nylon fishing line

**Figure 2.**
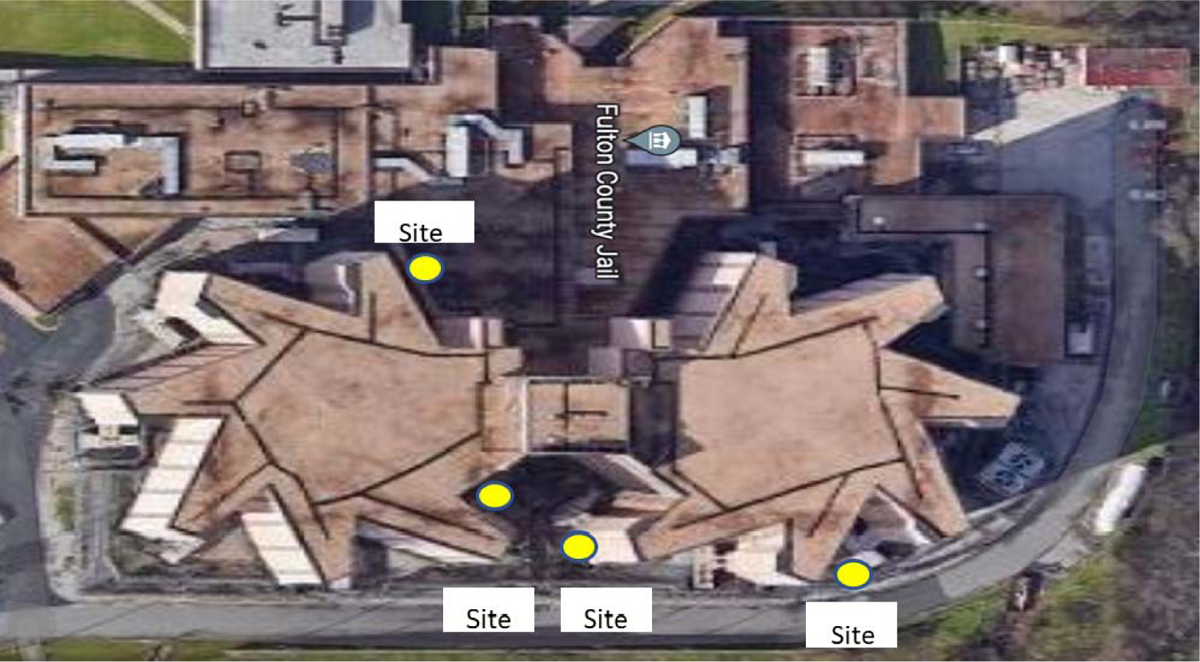
Manhole locations used for wastewater sample collection on the Fulton County Jail property (Google Maps)

For this study, we assumed that the source of wastewater flowing under each manhole corresponded to only the zone of the building in closest proximity to the collection point. This assumption was tested by adding tracer dye (EcoClean Solutions, Copiague NY) into toilets in on various housing blocks. When the dye from one housing block was detected in wastewater at multiple manholes, we assumed that the sewerage lines from multiple housing units were connected and that the wastewater at one manhole collection point could come from multiple housing blocks. For this analysis, we focused on wastewater data from a single, downstream collection point, Site 3 (See Figure 2), which contained a mix of wastewater from the south and north towers and used this as a proxy for the wastewater concentration of SARS-CoV-2 for the entire jail.

### COVID-19 Individual Testing: Rapid Test Collection

Healthcare staff offered universal opt-out rapid antigen testing to entrants and collected specimens at intake (BinaxNOW, Abbot Laboratories, Chicago, IL through 1/31/22; QuickVue, Quidel Corporation, San Diego CA starting 2/1/22). This diagnostic testing was part of the jail’s daily, standard entry protocol. Later in jail custody, antigen testing was available if residents exhibited COVID-19 symptoms, or if an individual made a request.

### COVID-19 Individual Testing: PCR Nasal Swab Collection

Concurrent with initiating WBS, a team from Emory University began a mass screening program in October 2021. We piloted self-collection of nasal specimens for PCR testing by the jail residents using a SteriPack swab [SteriPack, Lakeland FL], following a previously reported qualitative study of the acceptability of the novel specimen collection strategy [1]. On dates when mass collection of nasal swabs was performed, we hired off-duty correctional officers to escort the Emory specimen collection teams to areas of the jail to distribute and retrieve the nasal swabs. Bar codes, pre-printed on the tubes, were scanned into a cloud-based registration portal system for the diagnostic testing laboratory (Northwell Health Laboratory, Lake Success, NY) and minimized the time from specimen collection to registration to less than a minute per swab. We shipped swabs overnight to the laboratory for RT-PCR analysis via an LGC, Biosearch Technologies SARS-CoV-2 ultra-high-throughput End-Point RT-PCR Test (BT-SCV2-UHTP-EP) to detect positive nasal swabs (Biosearch Technologies, Hoddesdon, UK).

espite rapidspecim encollection, insufficient staffing precluded mass screening the entire resident population on any one occasion. Each week, areas of the jail screened by RT-PCR tests were either randomly selected or targeted, based on known ongoing outbreaks, to provide information for appropriate isolation and quarantine. We obtained nasal swabs for RT-PCR from a portion of the housed residents and used the results to develop a proxy measure for the prevalence of SARS-CoV-2 infection in the population. We from 2,497 to 2,904. The majority of residents in the jail during the study period were male and 88.8% were Black. There were 17 mass diagnostic PCR testing events in the jail which resulted in a total of 3,770 self-collected swabs tested by RT-PCR. Rapid COVID-19 diagnostic test results were collected every week (28 weeks) and totaled 9,975 over the study period. Weather and holiday schedules permitted weekly wastewater sampling on 25 collection days.

### Detection of SARS-CoV-2 in Jail Wastewater

In total, 79 samples were collected from 4 manholes sites (Figure 2). The Spearman’s correlation coefficients show strong correlations between Ct values of wastewater samples collected from different sites on the same day (Table S2, Figure S2), which indicates that results from one site could be used as a proxy for wastewater from the entire jail. The flow of wastewater was examined through sewage mapping via dye testing (REF). This showed that wastewater from both towers accumulated in site 3 and the results from this site were used for further analyses.

SARS-CoV-2 was detected in 80% of 25 Moore swab samples of wastewater collected from site 3 during the study period. Of the 20 samples that tested positive between 20 October 2021 and 4 May 2022, the mean Ct value was 33.94 (SD=3.74) and the median was 34.69. Samples were not collected during three holiday weeks in November-December 2021.

There was considerable temporal variability in the wastewater Ct values during the study period (Figure 3). The wastewater Ct value declined sharply from the sample collected during the week of 15 December 2021 to the sample collected during the week of 5 January 2022. The minimum Ct value for the whole study period (28.1, likely corresponding to the peak SARS-CoV-2 concentration in the jail wastewater) was measured in a sample from 5 January 2022 during the Omicron surge in Atlanta. The wastewater Ct values were in this range for five consecutive weeks (Figure 3). No SARS-CoV-2 RNA was detected in wastewater samples from one sampling date in November 2021 and four sampling dates in March-April 2022.

**Figure 3.**
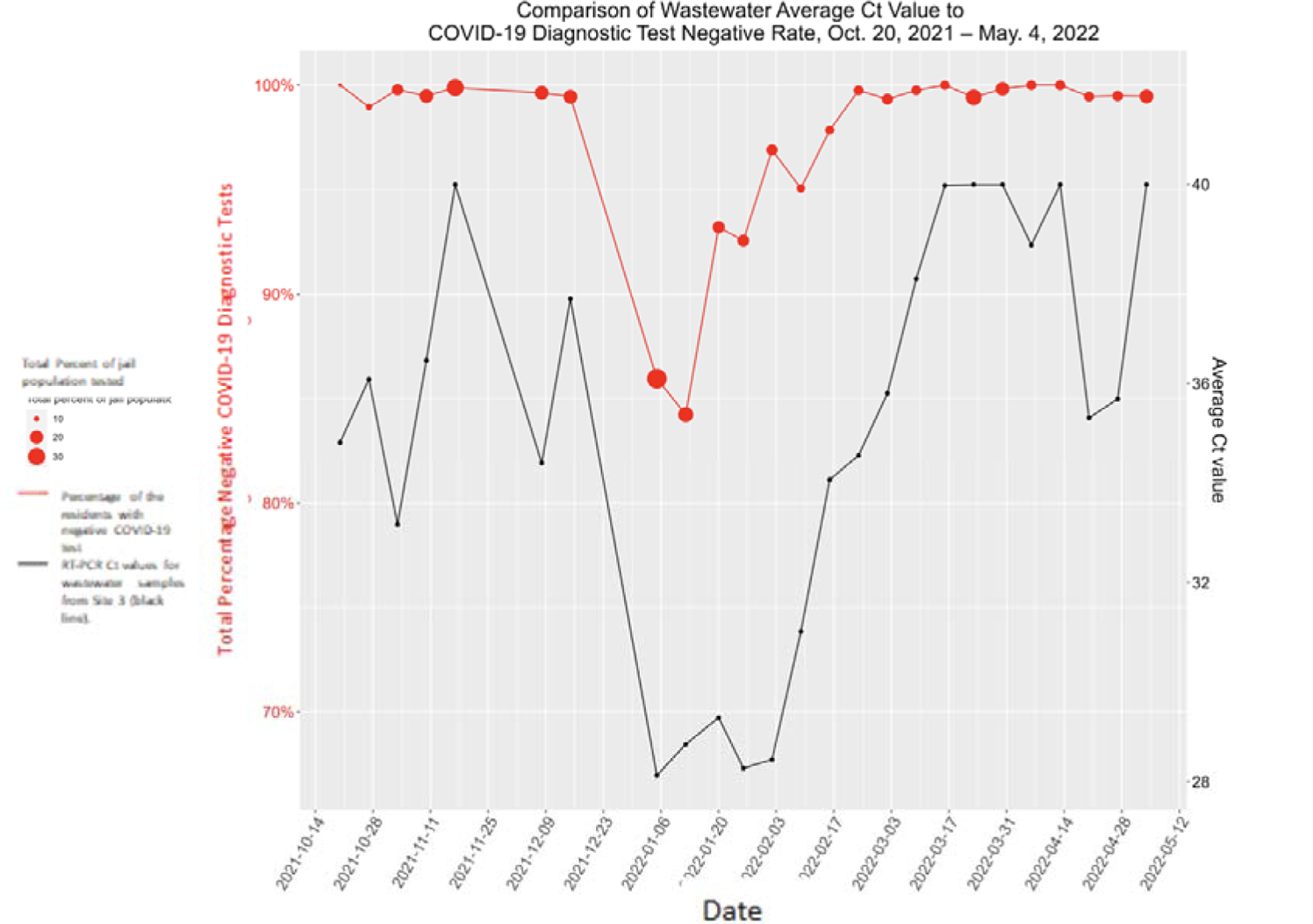
SARS-CoV-2 Ct Values for Wastewater and COVID-19 Diagnostic Results for Jail Residents by week, October 20, 2021 - May 4, 2022, Fulton County (GA) Jail.

### COVID-19 Diagnostic Testing

Table 2 summarizes COVID-19 diagnostic testing. The median number of total diagnostic tests conducted each week was 443 tests, and the majority of these were rapid antigen tests administered by the jail healthcare vendor. The median number of weekly rapid antigen tests was greater (363) than the number of PCR diagnostic tests that were administered during the mass testing events by the research study (mass testing event median = 186). The overall test positivity averaged 3.4% (SD= 6.6%) over the study period.

Figures 3 and 4 present the number of weekly COVID-19 diagnostic tests administered and the percentage of those that were positive over the study period. The total percentage of the jail population tested fluctuated but increased as the study progressed (Figure 3). Most was done via rapid antigen tests (Figure 4). Notable peaks in testing and test positivity occurred during the last weeks of December 2021 and early January 2022 during the Omicron surge in Atlanta.

**Figure 4.**
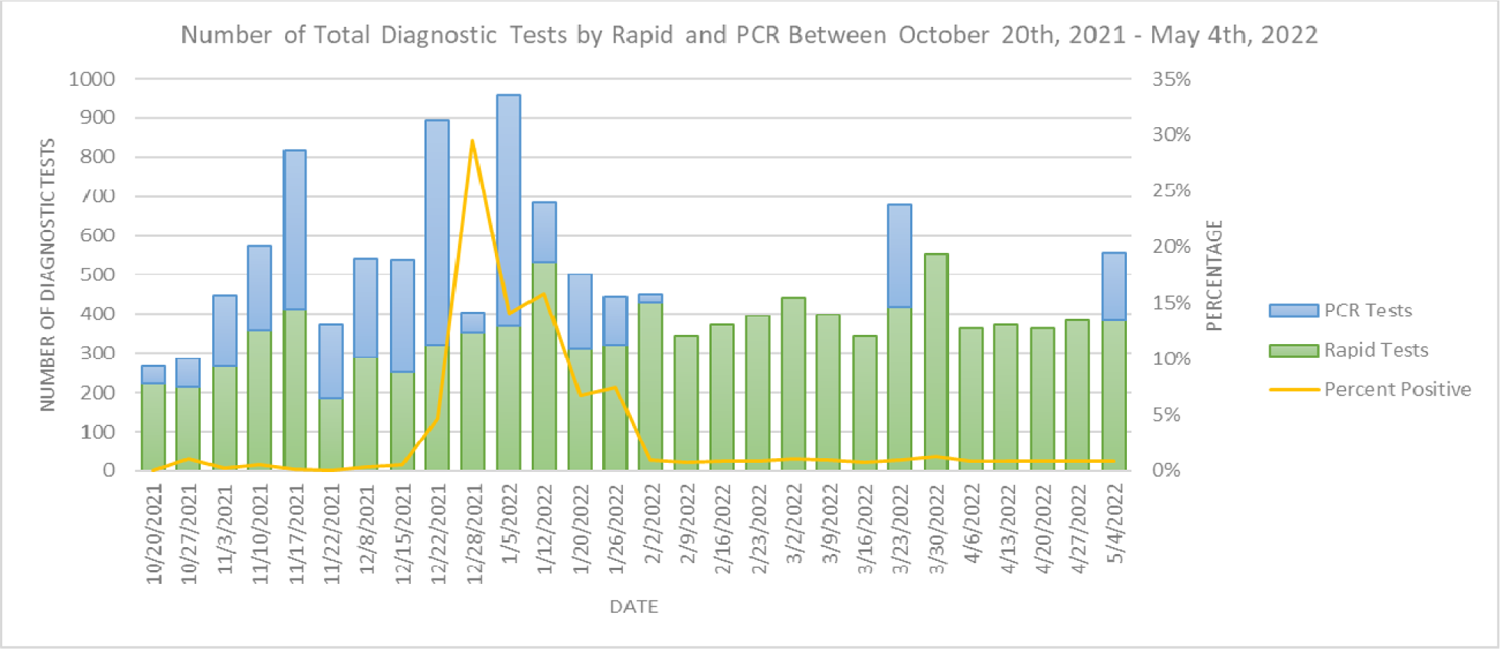
Total number and type of COVID-19 diagnostic tests of jail residents and percentage of positive tests by week, October 20, 2021 – May 4, 2022. Fulton County (GA) Jail.

the weekly test positivity rate correlated with the total percentage of the jail residents that were tested (rho=0.308, p-value=0.11). (Figure 3).

Overall, the PCR tests consistently had a higher positivity rate than the routine rapid antigen testing. During the midwinter surge, there was a much higher proportion of positive PCR tests (e.g., week of December 28, 2021, 63.5%) compared to the proportion of positive rapid antigen tests (24.4%). Nonetheless, the positivity rates for the PCR test and rapid antigen test were correlated during weeks when both tests were administered (r= 0.65, p-value=0.004).

### Relationship between COVID-19 Diagnostic Test Results and Wastewater Monitoring Results

The relationship between the COVID-19 diagnostic test results and WBS results were examined in several ways. When the diagnostic test positivity rate was low for several weeks, e.g., March 9, 2022 – April 13, 2022, the Ct values for the wastewater samples were high (38.1-40) or negative, indicating little or no detection of SARS-CoV-2 RNA in the wastewater samples. Conversely, low Ct values were measured in the wastewater samples during the weeks when the COVID-19 test positivity rates were high, e.g., early January 2022. Overall, the total COVID-19 diagnostic test positivity rate had a strong negative correlation with the wastewater Ct values combined over time (Spearman’s Correlation Coefficient: r= −0.67, p-value= <0.01) (Table 3).

All diagnostic tests were grouped into one datapoint representing each week between October 20, 2021 – May 4, 2022. Spearman correlation coefficients were calculated within the variable grouping of diagnostic test positivity rates and between variable groupings of diagnostic positivity rates and wastewater results (Table 3). Logistic regression was also used to analyze the relationship between the diagnostic positivity and the WBS results as a dichotomous outcome (presence/absence of SARS-CoV-2). Holding all other predictors constant, the odds of a positive WBS reading increased by 4.773 (95% CI 3.701-5.845) for a one unit increase in diagnostic test percent positivity (Figure S3).

## Discussion

prevalence of jail residents’ COVID-19 positive tests correlated with SARS-CoV-2 detection in the jail wastewater over time, which provides evidence that WBS can serve as an indicator of the jail population infection and its residents’ viral load. We also tested the feasibility and practicality of conducting both individual COVID-19 testing with self-collected nasal swabs and WBS on a routine basis. While gathered self-collected specimens rapidly, our inability to test all jail residents in a single shift supports the need for an aggregate measurement of population infection. Overall, our data indicate that WBS was a sensitive signal of when COVID-19 cases were present in the jail population and of surges in infection. [27, 28]

While the median number of rapid and PCR tests differed during the study period (363 and 186, respectively), the strong correlation between the positivity rate of the two different tests (r= 0.65, p-value= <0.01) suggests relatively accurate results from both forms of diagnostic tests and a predisposition to test hotspot areas.

During the fall of 2021, there was an upward trend in the portion of the jail population that participated in the mass testing events (Figure 3). This occurred by establishing an efficient screening method and having our testing routine improve over time so that each swab could be gathered and registered in under a minute. The jail’s housing configuration (cells rather than open dormitories) slowed the collection process. Nonetheless, we demonstrated that our process could achieve specimen collection from multiple housing units per hour. Ideally, if we could sample wastewater from specific housing units, it may be possible to identify the housing area that was the source of SARS-CoV-2 detected in the wastewater and use this information to focus diagnostic testing on a smaller portion of the jail. The current mass testing methods would then be adequate to test the subpopulation with the positive wastewater sample.

There was one defined peak of diagnostic test positivity over the duration of the study on December 28, 2021 of 29.5% (Figure 3). The spike in COVID-19 cases in the jail (January 5, 2022) occurred 8 days prior to a community surge in Fulton County and aligned with COVID-19 case surges in Atlanta and nationwide due to the Omicron variant [29]. Therefore, jails may serve as an early warning signal for community spikes for COVID-19 and other infectious diseases detectable in wastewater.

This study demonstrated the efficiency and feasibility of conducting WBS for SARS-CoV-2 on a regular basis in a jail setting. Other jails have been conducting wastewater studies, but to our knowledge have yet to report a correlation with the proportion of the jail population testing positive for COVID-19 [30, 31]. WBS has been studied in non-carceral institutions, such as university residence halls, where the turnover rate of who resides in the dorm is low and students normally live an entire semester in the same room, but can have frequent visitors [16-18, 20, 32, 33]. In contrast, the median length of stay for a Fulton County Jail resident has historically been 5 days and the mean stay 22 days.[22]. This contrast between residence times in university dormitories versus a jail may account for different utility of WBS data in this jail-based study compared to other settings.

Similar to the experience of WBS studies at university campuses, the collection and processing of Moore Swab samples from the jail are much less expensive and faster than individual diagnostic testing of the jail residents [32]. A report on costs of WBS in this study is pending, but they compare favorably with the previously reported WBS study of university residence halls, where ten Moore Swabs cost $12, sample collection time averaged 30 minutes, sample processing time was 5 hours, and the overall turnaround time from sample collection to reporting final lab results was 2 to 3 days [18, 32].

### Strengths

The primary objective of this study was to examine if WBS is an effective and feasible strategy to monitor for COVID-19 outbreaks in a carceral setting. Strengths of this study are:

- Large jail population allowed for a reduction of diagnostic testing sampling bias.
- Weekly iterations of the mass diagnostic testing procedure allowed for several opportunities to enhance the number of diagnostic test results.
- High number of weekly diagnostic tests enabled us to compare the positivity rates with the WBS data.
- Weekly collection and analysis of WBS samples in tandem with the diagnostic tests.
- We eliminated the human error in data entry when we eliminated typing test identification barcodes into the portal and instead began scanning them straight into the portal.
- Close collaboration with jail officials provided the exclusive opportunity to conduct a prolonged study (6 months) within the jail, capturing the entirety of the Omicron variant peak.

### Limitations

The current study had several limitations:

- Jail size, availability of escorts, and limited resources for testing team precluded diagnostic testing every week, or the entire jail population in any one week; results from the portion of the jail population that was tested for COVID-19 was used as a proxy for COVID-19 prevalence in the entire jail population.
- The data were derived from testing for public health practice, and individual testing was not conducted on a random basis but rather for individuals as needed for the purposes of an infection control program. Nonetheless, on no week was testing confined only to areas of the jail known to have high COVID-19 prevalence. For many weeks, rapid antigen tests were mainly administered to new residents at intake rather than the general jail population, and PCR testing was targeted to other areas in the jail.
- The RT-qPCR results (Ct values) for the Moore swab samples should be considered semi-quantitative. The concentration of SARS-CoV-2 RNA per volume of wastewater is not easily calculated since the wastewater flow through the Moore swab during the time it was submerged in the manhole is not known.
- The Ct values from the Moore swab samples provide only an approximate indication of the presence and number of COVID-19 cases in the jail due to uncertainty about the

magnitude and duration of SARS-CoV-2 fecal shedding and unknown dilution from the volume of wastewater [18]. Nonetheless, the strong correlation between the Ct Values and COVID-19 individual test positivity rate suggests that WBS is an effective method to surveil for SARS-CoV-2 in a jail setting.

- A jail is not a closed system: many people are entering and leaving on a daily basis. A resident who sheds fecal matter containing SARS-CoV-2 on one day may leave the jail before the next round of individual screening for COVID-19.
- Only COVID-19 tests of residents were included in our analyses, but staff contribute to wastewater. Since residents greatly outnumbered staff, fecal material from staff probably had a negligible effect on the WBS results. Jail staff were periodically offered diagnostic testing at roll call, and their test positivity patterns mirrored the trends for the residents (data not shown).
- Weekly COVID-19 test results were aggregated from Sunday to Saturday, whereas wastewater sample collection occurred mid-week. We might have observed a stronger correlation between the wastewater and diagnostic test results if the wastewater samples were collected more frequently.

## Conclusion

Even under ideal circumstances--external funding, collegial working relationships, and adequate resources--administering individual COVID-19 diagnostic tests to the entire Fulton County Jail on a weekly basis was not a feasible COVID-19 surveillance strategy. However, conducting WBS at the jail was highly effective and sustainable. The WBS results aligned well with the COVID-19 diagnostic test positivity rates among jail residents and could serve as a sensitive and economical surveillance tool for COVID-19 in a large jail facility. Overall, the high resident turnover rate in the jail, the countywide geographic range of the subjects entering the jail, and the high proportion of wastewater samples with SARS-CoV-2 detection suggest that WBS at the jail is important not only for understanding COVID-19 burden in the jail and using the results to guide public health response to mitigate transmission but also that jails could serve as a valuable sentinel site for monitoring trends in COVID-19 prevalence and genetic variants in the wider community.

## Data Availability

All data produced in the present study are available upon reasonable request to the authors

## Acknowledgments

We wish to thank the Fulton County Sheriff Patrick Labat and the Sheriff’s Office, Fulton County Jail staff, and the FCJ team members of NaphCare and Johnson Controls, whose help has been invaluable. We are grateful for the residents of FCJ who participated in this public health project. We thank Jamie VanTassell and Lauren Briggman for wastewater sample collection; Orlando Sablon, Lizheng Guo, Matthew Cavallo, Caleb Cantrell, Jillian Dunbar for laboratory analyses; and Haisu Zhang, Lutfe-E-Noor Rahman, and Weiding Fang for wastewater data management and analyses.

