## Supplementary figures and images for "Wastewater Surveillance for SARS-CoV-2 in an Atlanta, Georgia Jail: A study of the feasibility of wastewater monitoring and correlation of building wastewater and individual testing results"

### Figure S1

Figure S1

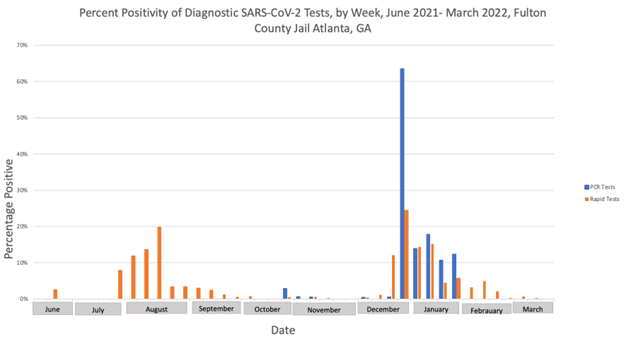

### Figure S2

Figure S2. Average Ct Values over time by manhole, four separate locations, October 2021-May 2022.

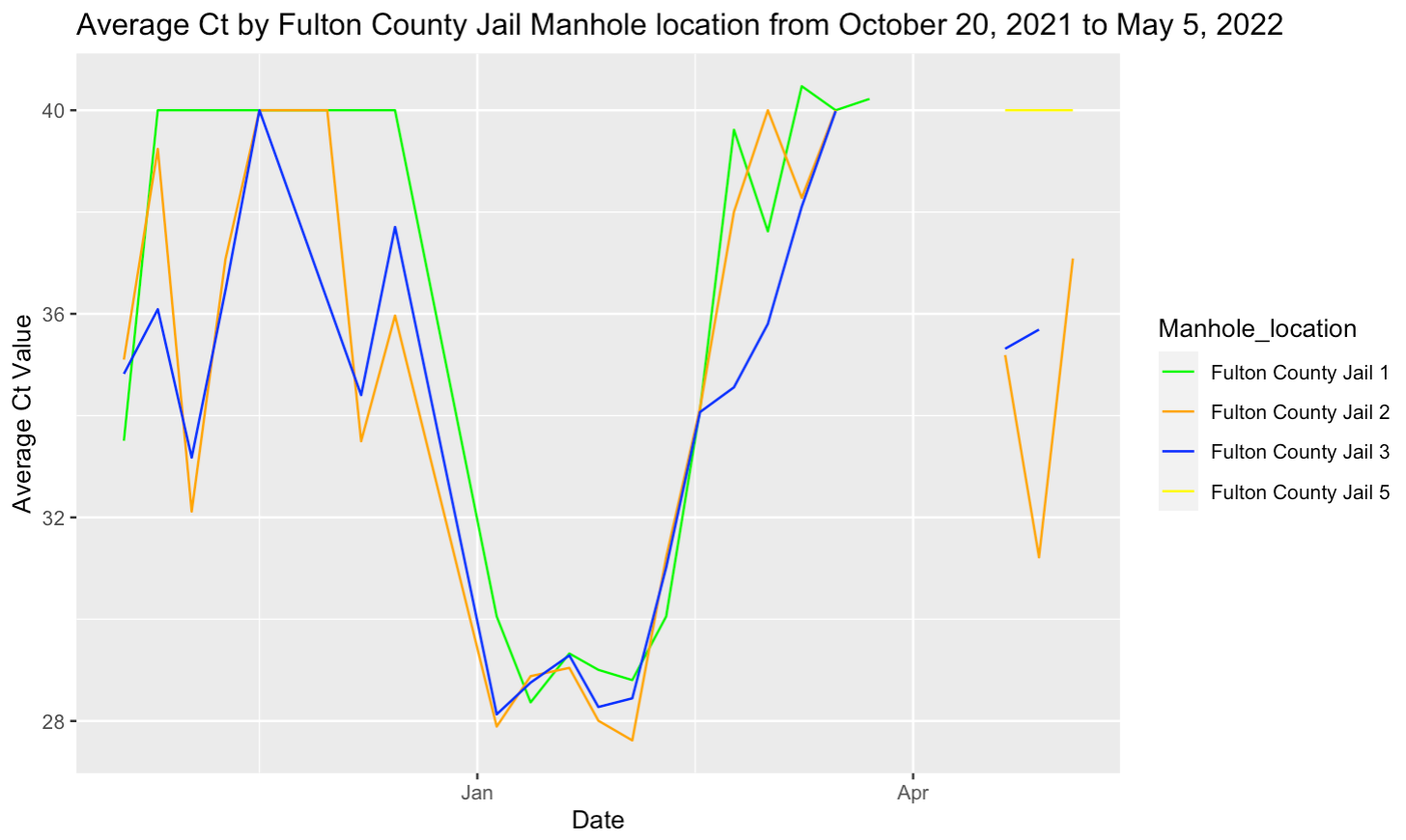
