## Supplementary material for "Wastewater Surveillance for SARS-CoV-2 in an Atlanta, Georgia Jail: A study of the feasibility of wastewater monitoring and correlation of building wastewater and individual testing results": Figure S3

Figure S3. Logistic regression of diagnostic test percent positivity and wastewater as a dichotomous outcome (positive or negative), October 2021-May 2022.

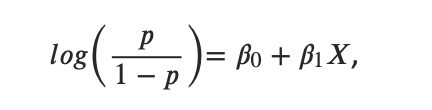

Where p is the probability of detecting SARS-CoV-2 in the wastewater sample from site 3 and X is the percent of positive COVID-19 diagnostic tests. The estimates of *𝛽0* and*𝛽1* were 0.484 and 4.773, respectively.
