## Supplementary material for "Wastewater Surveillance for SARS-CoV-2 in an Atlanta, Georgia Jail: A study of the feasibility of wastewater monitoring and correlation of building wastewater and individual testing results": Figure S4

Figure S4. SARS-CoV-2 wastewater results by dichotomous outcome and COVID-19 diagnostic test positivity rates October 2021-May 2022.

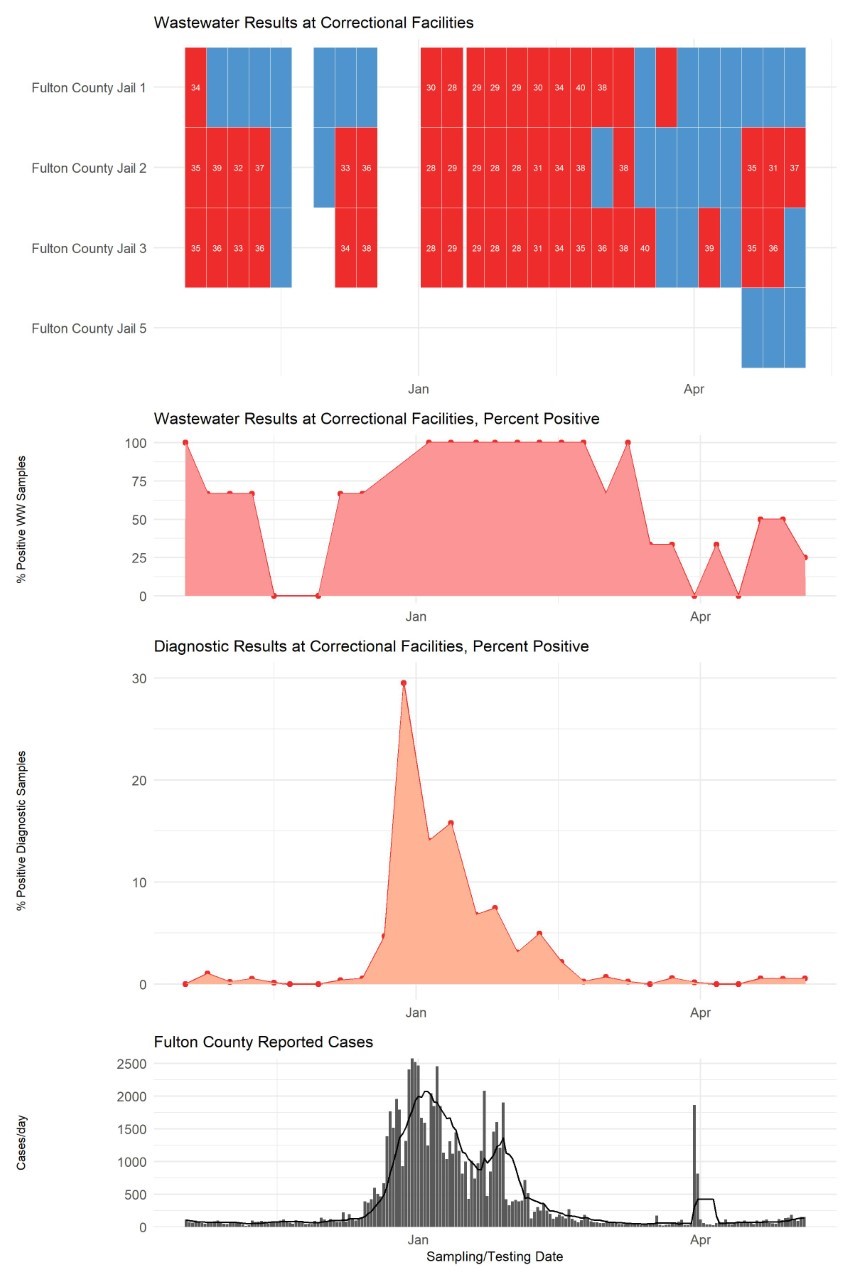
