## Supplementary material for "Wastewater Surveillance for SARS-CoV-2 in an Atlanta, Georgia Jail: A study of the feasibility of wastewater monitoring and correlation of building wastewater and individual testing results": Table S1

Table S1. Of the tests administered in Table S1, those that resulted as positive by week and type of test, Fulton County Jail, October 5, 2021 – May 4, 2022.

| Descriptions of variables used for analysis | |
| --- | --- |
| Variable | Description |
| PCR Tests | Those tests which the SARS-CoV-2 virus was detected. |
| Jail Population | The total resident population of the Fulton County Jail determined through the resident count on weeks that the study team received rosters from jail personnel (point count of the jail). On the weeks that no rosters were received, public records from the Georgia Department of Community Affairs (DCA) were used to estimate the jail population (Georgia Department of Community Affairs, 2022). |
| Positivity Rate | The total number positive diagnostic tests (PCR + rapid) in a week divided by the total number of tests administered in the same week. |
| Percent of Jail Tested | The total number of diagnostic tests (PCR + rapid) administered in a week divided by the jail population in the same week. |
| Ct Value | RT-qPCR cycle threshold value measured in wastewater samples at the Emory laboratory using primers and probes for the N1 gene of SARS-CoV-2 |
