## Supplementary material for "Wastewater Surveillance for SARS-CoV-2 in an Atlanta, Georgia Jail: A study of the feasibility of wastewater monitoring and correlation of building wastewater and individual testing results": Table S2

Table S2. Spearman’s correlation coefficients of all Ct Value wastewater results between manhole sites 1-4. All values were statistically significant (alpha < 0.05), October 2021-May 2022.

| **Ct Values Correlated between Manhole Sites (1-4)** | | | | |
| --- | --- | --- | --- | --- |
|  | Manhole 1 | Manhole 2 | Manhole 3 | Manhole 5 |
| Manhole 1 | 1 |  |  |  |
| Manhole 2 | 0.7437932 | 1 |  |  |
| Manhole 3 | 0.8324621 | 0.8945793 | 1 |  |
| Manhole 5 | 0.7075317 | 0.6935212 | 0.8041958 | 1 |
