## Supplementary material for "Wastewater Surveillance for SARS-CoV-2 in an Atlanta, Georgia Jail: A study of the feasibility of wastewater monitoring and correlation of building wastewater and individual testing results": Table 1

Table 1. Demographic characteristics of residents in the Fulton County Jail, Main Complex, October 20, 2021 to May 4, 2022.

| *Reported Sexual Assignment* | *Percent of Population* |
| --- | --- |
| Male | 98.4 |
| Female | 1.6 |
| *Race/ Ethnicity* |  |
| Black, non- Hispanic | 88.8 |
| White, non- Hispanic | 10.3 |
| Hispanic | <1 |
| Other | <1 |
| *Charges* |  |
| Misdemeanor only | 6.8% |
| Felony | 93.2% |

*Source: Fulton County Jail.*
