## Supplementary material for "Wastewater Surveillance for SARS-CoV-2 in an Atlanta, Georgia Jail: A study of the feasibility of wastewater monitoring and correlation of building wastewater and individual testing results": Table 2

Table 2. Summary of COVID-19 diagnostic testing results, October 2021-May 2022, Fulton County Jail, Atlanta, GA

| ***COVID-19 Diagnostics Weekly*** | | *Mean (SD) per week* | *Median per week* | *Min, Max per week* | *Totals over entire study* |
| --- | --- | --- | --- | --- | --- |
|  | All Diagnostic Tests | 491 (176) | 443 | 267, 961 | 13,745 |
|  | Rapid Tests | 356 (84) | 363 | 186, 554 | 9,975 |
|  | PCR Tests | 222 (167) | 186 | 20, 591 | 3,770 |
|  | % of Jail Tested^a^ | 18.3 (7.1) | 16 | 9.7, 38.2 |  |
|  | Overall Test Positivity Rate^b^ | 3.39 (6.56) | 0.55 | 0, 29.5 |  |
| a Numerator is the number of positive tests in a given week, denominator is the jail population for the week  b Numerator is the number of positive tests in a given week, denominator is the total tests for the same week | | | | | |
