## Supplementary material for "Wastewater Surveillance for SARS-CoV-2 in an Atlanta, Georgia Jail: A study of the feasibility of wastewater monitoring and correlation of building wastewater and individual testing results": Table 3

Table 3. Spearman correlation coefficients for wastewater Ct values and diagnostic test positivity rates compared within their variable groupings and between variable groupings. Each datapoint is correlated with all other datapoints, none are grouped based on date or other variables. **Fulton County Jail, GA October 20, 2021-May 4, 2022.**

| **Spearman Correlation Coefficients for Wastewater and Diagnostic Test results** | | | |
| --- | --- | --- | --- |
| **Diagnostic Tests** | % Positive PCR^1^  R (p-value) | % Positive Rapid Antigen^2^  R (p-value) | Total % Positive Tests^3^  R (p-value) |
| % Positive PCR | 1.00 | 0.91 (<0.01) | 0.78 (<0.01) |
| % Positive Rapid Antigen |  | 1.00 | 0.97 (<0.01) |
| Total % Positive Tests |  |  | 1.00 |
| **Wastewater and Diagnostic Correlation** | % Positive PCR | % Positive Rapid | Total % Positive Tests |
| Wastewater Ct Values | -0.54 (0.048) | -0.64 (<0.01) | -0.67 (<0.01) |
| ^1^ Number of positive PCR tests in one week over the total number of PCR tests administered for the same week  ^2^ Number of positive rapid tests in one week over the total number of rapid tests administered for the same week  ^3^ Total number of positive diagnostic tests (PCR + rapid) in one week over the total number of diagnostic tests administered for the same week | | | |
